# Somatic mutation of *ELF4* causes autoinflammatory diseases and cell type-specific immune alterations

**DOI:** 10.64898/2026.04.08.26350315

**Authors:** Qianlu Zhang, Yatang Lei, Xiaodong Zhao, Hongqiang Du

**Affiliations:** Department of Rheumatology & Immunology Children’s Hospital of Chongqing Medical University, National Clinical Research Center for Child Health and Disorders, Ministry of Education Key Laboratory of Child Development and Disorders, Chongqing 400014, China; Chongqing Key Laboratory of Child Rare Diseases in Infection and Immunity, Chongqing 400014, China; Jiangxi Children’s Medical Center, Jiangxi Hospital Affiliated to Children’s Hospital of Chongqing Medical University, Jiangxi Maternal and Child Health Hospital, Nanchang, Jiangxi 330006, China

**Author notes:** **Correspondence Corresponding authors Hongqiang Du**, Department of Rheumatology & Immunology Children’s Hospital of Chongqing Medical University, Chongqing 400014, China, E-mail address, **Xiaodong Zhao**, Department of Rheumatology & Immunology Children’s Hospital of Chongqing Medical University, Chongqing 400014, China. These authors contributed equally: Qianlu Zhang, Yatang Lei.

**Keywords:** ELF4, Somatic mutation, autoinflammatory diseases, Inborn errors of immunity

## Abstract

ELF4 is an ETS family transcription factor involved in immune regulation, and germline loss-of-function mutations in ELF4 have been known as “deficiency in ELF4, X-linked”(DEX). To date, ELF4-related disease has been exclusively associated with germline mutations. Here, we report a pediatric patient with recurrent mucocutaneous inflammation and periodic fever caused by a somatic truncating mutation in ELF4. By directly comparing ELF4-mutant and wild-type immune cells within the same individual using full-length single-cell RNA sequencing, we identified mutation-associated transcriptional alterations across multiple immune cell types. Pathway analyses revealed cell type-specific immune alterations, characterized by reduced antiviral and interferon-related signaling in NK cells and enhanced inflammatory pathways related to Th17 differentiation and inflammatory bowel disease in CD16^+^ monocytes. This study expands the disease spectrum of ELF4 deficiency by identifying somatic truncation of ELF4 as a genetic mechanism underlying autoinflammatory diseases and biased immune programs.

## Introduction

ELF4 is a member of the ETS family of transcription factors^1^ and plays an important role in immune regulation, including antiviral responses and inflammatory signaling^2^. Germline loss-of-function mutations in ELF4 have been identified as the genetic cause of deficiency in ELF4, X-linked (DEX), a disorder characterized by recurrent mucocutaneous inflammation, systemic inflammatory manifestations, and variable immune abnormalities^3,4^. Clinical presentations of DEX are heterogeneous, ranging from isolated mucosal ulcers to severe inflammatory bowel disease-like phenotypes, highlighting the context-dependent role of ELF4 in immune homeostasis. To date, ELF4-related disease has been exclusively associated with germline mutations^3–8^.

Somatic mutations arising in hematopoietic cells have emerged as important contributors to immune-mediated diseases^9^. Comparing mutant and wild-type immune cells within the same individual provides a powerful framework for dissecting mutation-associated immune programs while minimizing inter-individual variability. In this context, full-length single-cell RNA sequencing has become a valuable approach for resolving cell type–specific transcriptional alterations associated with somatic mutations^10^. Here, we report a pediatric patient harboring a somatic truncating mutation in ELF4 and combine genetic, functional, and single-cell transcriptomic analyses to delineate the immune alterations associated with somatic ELF4 deficiency.

## Materials and methods

### Patient

The ethics committee of Children’s Hospital of Chongqing Medical University approved the study. Written informed consent for participation in the study was obtained from patient’s parents. Blood samples from patient and family members were collected for genetic studies, which were performed in accordance with the Declaration of Helsinki.

### Genetic Analyses

Genomic DNA were isolated from peripheral blood samples and buccal mucosa. Disease-causing mutations were screened using whole genome sequencing (WES) (Fujun Genetics, Inc.). Candidate mutations were confirmed by Sanger sequencing using the following primers: seq F:5’-CATAGCTGGAAGACCCCTC-3’, seq R:5’-CCCCTCCTTACCTAACTCTCC-3’.

### Construction of plasmids

The 7.1-pCMV-3×Flag expression vectors encoding wild-type human ELF4 were generated as previously described^7^. Point mutations were introduced into the wild-type constructs by overlap-extension PCR, and the resulting plasmids were confirmed by Sanger sequencing.

### Cells

HEK 293T (Human Embryonic Kidney 293 cells transformed by expression of the large T antigen from SV40) (ATCC, CRL-11268) were cultured in Dulbecco’s modified Eagle medium (DMEM).

### Full-length single-cell RNA sequencing

Full-length single-cell RNA sequencing (SeekOne ® DD scFAST-seq^11^) was performed on peripheral blood mononuclear cells (PBMCs) obtained from the patient harboring a somatic truncating mutation in ELF4. Fresh PBMCs were isolated and processed according to standard procedures to ensure high cell viability prior to library preparation. Single-cell RNA-seq libraries were constructed using the SeekOne® Single Cell Whole Transcriptome Kit (SeekGene) following the manufacturer’s instructions, loaded onto the SeekOne® DD Chip S3, and sequenced on an Illumina NovaSeq 6000 platform with paired-end 150 bp reads.

### Preprocessing, quality control, and cell clustering

Raw sequencing data were processed using fastp to remove adaptor sequences and low-quality reads, followed by analysis with the SeekSoul pipeline for demultiplexing cell barcodes and unique molecular identifiers (UMIs) and alignment to the human reference genome (GRCh38). Gene expression matrices were generated based on UMI counts. Cells with low UMI counts or excessive mitochondrial gene expression were excluded to ensure data quality. Downstream analyses were performed using the Seurat R package. Gene expression data were normalized, highly variable genes were identified, and principal component analysis (PCA) was conducted. The first 15 principal components were used for unsupervised clustering, and Uniform Manifold Approximation and Projection (UMAP) was applied for dimensionality reduction and visualization of major immune cell populations.

### Cell type annotation and compositional analysis

Cell clusters were annotated based on the expression of canonical marker genes, enabling reliable identification of major immune cell populations, including T cells, B cells, NK cells, CD14+monocytes, CD16+monocytes, and neutrophils. ELF4-mutant and wild-type cells were overlaid onto the UMAP representation to assess their distribution across immune compartments. The relative proportions of mutant and wild-type cells within each immune cell subset were compared to evaluate differences in cell type composition.

### Differential gene expression analysis

Differential gene expression analyses were performed within each immune cell subset by comparing ELF4-mutant cells with wild-type cells from the same patient using the FindMarkers function in Seurat. Genes with a P value < 0.05 and an absolute log2 average fold change > 0.1 were considered differentially expressed. Heatmaps and network visualizations were generated using the ggplot2 and heatmap2 packages in R.

### Single-cell mutation identification and analysis

Single-cell mutation analysis was performed on aligned sequencing data generated using Cell Ranger. Single-nucleotide variants were identified at the single-cell level using the Pysam-based tool cb_sniffer with default parameters. Reads lacking Chromium cellular barcode (CB) or unique molecular identifier (UMI; UB) tags were excluded^12,13^. For each UMI at a given genomic position, base calls were aggregated, and a mutation was assigned when a single base accounted for at least 75% of supporting reads. UMIs not meeting this criterion were excluded from downstream analyses.

### Pathway enrichment analyses

To interpret the biological significance of transcriptional differences between ELF4-mutant and wild-type cells, Gene Ontology (GO) biological process and Kyoto Encyclopedia of Genes and Genomes (KEGG) pathway enrichment analyses were performed on differentially expressed genes within each immune cell subset using the clusterProfiler R package. Pathways and GO terms with adjusted P values < 0.05 were considered significantly enriched.

### Statistical analysis

Samples were compared using two-tailed, unpaired Student’s t-test with GraphPad Prism 7.00. Error bars were represented by SEM. *P < 0.05, **P < 0.01, ***P < 0.001.

## Results

### Clinical manifestations of the patient

The proband (II.1) is a male patient who has experienced recurrent oral ulcers since the age of childhood. He developed recurrent episodes of fever, each lasting 1–3 days and occurring approximately every two weeks. No genital, perianal, or ocular ulcers were observed during the disease course. Other clinical features included normal growth and development, with no hepatosplenomegaly, lymphadenopathy, arthritis, or skin lesions noted.

Laboratory investigations revealed elevated inflammatory markers, with an increased C-reactive protein level of 20.02 mg/L (normal range: 0–10 mg/L), while the erythrocyte sedimentation rate remained within normal limits. Complete blood counts were unremarkable, and immunological evaluation showed no abnormalities. Following treatment with thalidomide, the inflammatory activity was effectively controlled.

### Genetic studies reveal a somatic nonsense mutation in *ELF4*

Based on the patient’s clinical and laboratory features suggestive of a monogenic autoinflammatory disorder, whole-exome sequencing was performed. Genetic analysis identified a *de novo* truncating variant in *ELF4* (c.239T>A, p.L80X) in peripheral blood cells from individual II.1 (Fig. 1A), which was absent in both parents. Furthermore, the mutation was not detected in the proband’s buccal mucosa tissue, supporting its classification as a postzygotic somatic mosaic mutation (Fig. 1B). The mutation introduces a premature stop codon in the N-terminal region of ELF4, predicting a severely truncated protein lacking critical functional domains. These findings establish that the proband carries a somatic truncating mutation in ELF4, providing genetic evidence for a somatic form of ELF4-related autoinflammatory disease.

**Figure 1.**
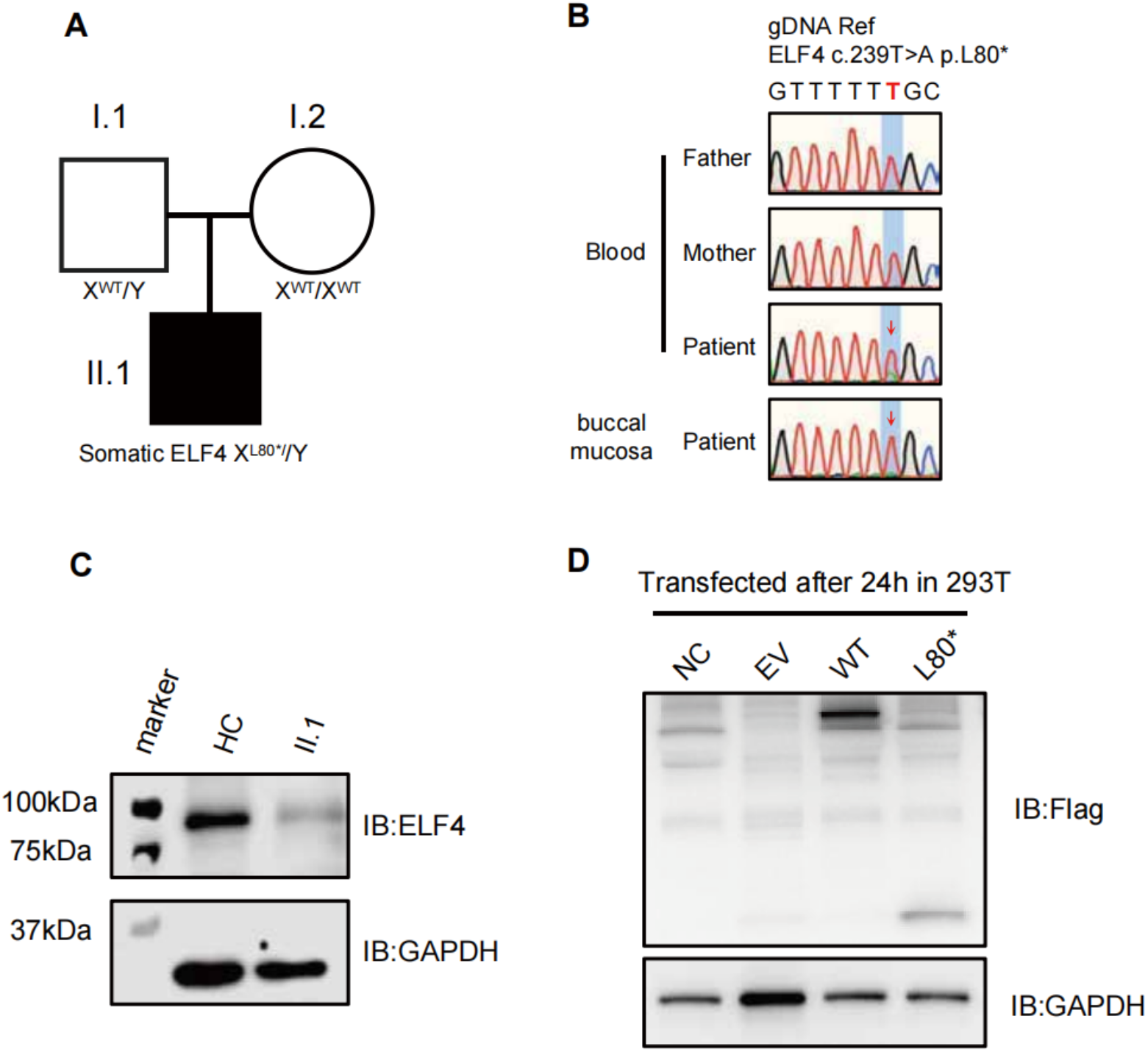
Identification and functional characterization of a somatic *ELF4* mutation. **(A)** Pedigree of the family showing the affected male proband (II.1) harboring a somatic truncating mutation in *ELF4* (p.L80X), while both parents are unaffected. **(B)** Sanger sequencing chromatograms showing the *ELF4* c.239T>A (p.L80X) variant detected in peripheral blood cells of the patient but absent in parental peripheral blood cells and buccal mucosa, supporting a postzygotic somatic mosaic mutation. **(C)** Immunoblot analysis of endogenous ELF4 protein expression in PBMCs from a healthy control (HC) and the patient (II.1). **(D)** Immunoblot analysis of FLAG-tagged wild-type (WT) and L80X-mutant ELF4 expressed in HEK293T cells.

### The L80X variant of ELF4 shows loss-of-function effects

To evaluate the functional impact of the p.L80X variant, we first examined endogenous ELF4 expression. Immunoblot analysis showed markedly reduced ELF4 protein in patient-derived cells compared with healthy controls, consistent with loss of function (Fig. 1C). We then transfected HEK293T cells with FLAG-tagged wild-type or L80X mutant ELF4 constructs. While wild-type ELF4 was robustly expressed, the L80X mutant protein was barely detectable, confirming its destabilizing effect (Fig. 1D).

Together, these data demonstrate that the p.L80X variant results in a clear loss of function. In contrast to germline *ELF4* deficiency, the somatic mosaic nature of this mutation likely restricts the proportion of affected immune cells, which may explain the relatively limited clinical phenotype in this individual. This distinction is summarized in Table 1.

**Table 1.** Comparison of clinical characteristics between individual II.1 and previously reported patients with germline ELF4 deficiency (DEX)

| Clinical feature | Individual II.1 (this study) | Reported DEX patients (germline ELF4 LOF)* |
| --- | --- | --- |
| Sex | Male | Predominantly male |
| Genetic status | Somatic mosaic ELF4 truncation (c.239T>A, p.L80*) | Germline hemizygous ELF4 loss-of-function variants |
| Mode of inheritance | Not inherited; parents negative | X-linked (inherited or de novo) |
| Age at onset | Childhood | Early childhood to adolescence |
| Oral ulcers | Present, recurrent | Common |
| Genital / perianal ulcers | Absent | Variable |
| Ocular involvement | Absent | Rare |
| Recurrent fever | Present | Common |
| Gastrointestinal involvement | Absent | Common (IBD-like colitis) |
| Skin manifestations | Absent | Variable |
| Arthritis / arthralgia | Absent | Variable |
| Inflammatory markers | Elevated CRP; ESR normal | Elevated CRP and/or ESR |
| Autoantibodies | ANA and anti-dsDNA negative | Typically negative |
| Lymphocyte subsets | Normal | Variable |
| Humoral immunity | Normal | Variable |
| Treatment | Thalidomide (effective) | Steroids, biologics (anti-IL-1, anti-TNF, anti-IL-12/23), others |
| Clinical course | Well controlled | Chronic, relapsing |

### Full length single-cell transcriptomic analysis reveals transcriptional alterations in ELF4-mutant immune cells

To characterize the transcriptional landscape associated with the somatic ELF4 mutation at single-cell resolution, we performed full-length single-cell RNA sequencing on peripheral blood mononuclear cells from individual II.1. Full-length scRNA-seq provides superior transcript coverage across the entire RNA molecule, allowing for detection of alternative splicing, isoform diversity, and gene fusions. It offers higher sensitivity per cell for low-abundance transcripts and accurate allele-specific expression analysis compared to 3’-end sequencing methods^14^. Unsupervised clustering identified all major immune populations—T cells, B cells, NK cells, CD14^+^ monocytes, CD16^+^ monocytes, and neutrophils—confirming robust cell-type resolution (Fig. 2A).

**Figure 2.**
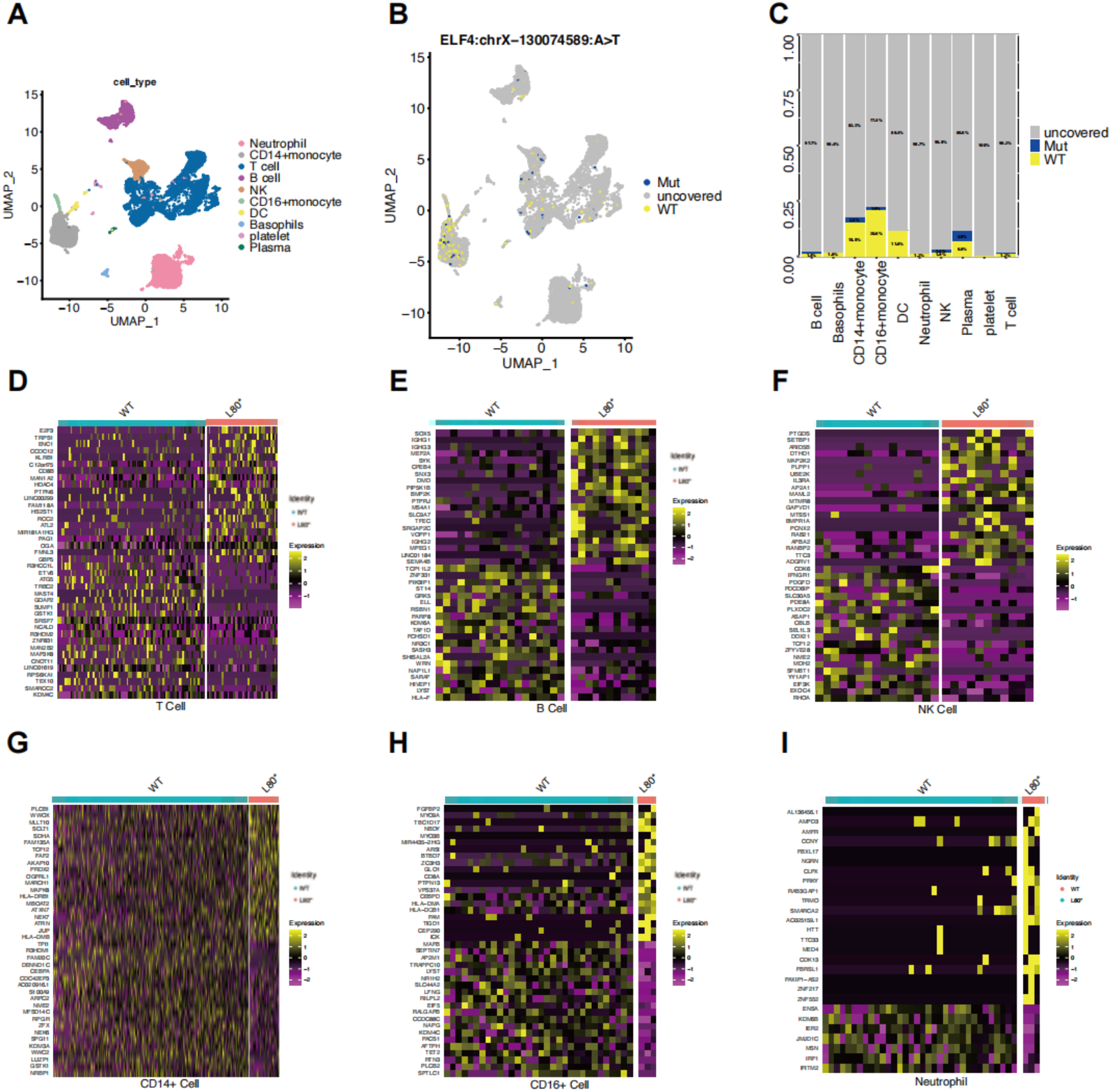
Single-cell analysis reveals cell type–specific immune consequences. **(A)** UMAP visualization of single-cell RNA sequencing data from patient PBMCs, colored by major immune cell types, including T cells, B cells, NK cells, CD14+monocytes, CD16^+^ monocytes, neutrophils, dendritic cells, basophils, platelets, and plasma cells. **(B)** UMAP projection highlighting the distribution of mutant ELF4 (L80*), wild-type (WT), and uncovered cells across immune cell populations, demonstrating mosaic representation of mutant cells in multiple lineages. **(C)** Proportion of mutant ELF4 (L80*) and wild-type cells within each immune cell population, showing variable lineage-specific representation of mutant cells. **(D-F)** Heatmaps showing differentially expressed genes between mutant ELF4 (L80*) and wild-type cells within lymphoid populations, including T cells, B cells, and NK cells. **(G-I)** Heatmaps showing differentially expressed genes between mutant ELF4 (L80*) and wild-type cells within myeloid populations, including CD14^+^ monocytes, CD16^+^ monocytes, and neutrophils.

Projection of ELF4-mutant and wild-type cells onto the UMAP representation showed that mutant cells were distributed across multiple immune lineages, albeit with variable frequencies (Fig. 2B). Accordingly, comparative analysis revealed differences in the proportion of mutant cells among subsets (Fig. 2C).

We next performed differential gene expression analysis within each immune subset. In lymphoid lineages (T, B, and NK cells), ELF4-mutant cells displayed distinct transcriptional profiles compared with their wild-type counterparts (Fig. 2D–F). Similarly, in myeloid subsets (CD14^+^ monocytes, CD16^+^ monocytes, and neutrophils), mutant cells also exhibited significantly altered gene expression patterns (Fig. 2G–I). These findings indicate that ELF4-mutant immune cells harbor mutation-associated transcriptional signatures across both lymphoid and myeloid compartments.

### Cell type–specific pathway alterations associated with somatic ELF4 mutation

To decipher the biological relevance of these transcriptional changes, we performed Gene Ontology (GO) and KEGG pathway enrichment analyses on differentially expressed genes within each immune subset.

In lymphoid cells, especially T and B cells, enriched pathways were related to immune activation and lymphocyte signaling—including antigen processing and presentation, T-cell receptor signaling, and B-cell receptor pathways—suggesting that the *ELF4* mutation perturbs adaptive immune regulation (Fig. 3A-D).

**Figure 3.**
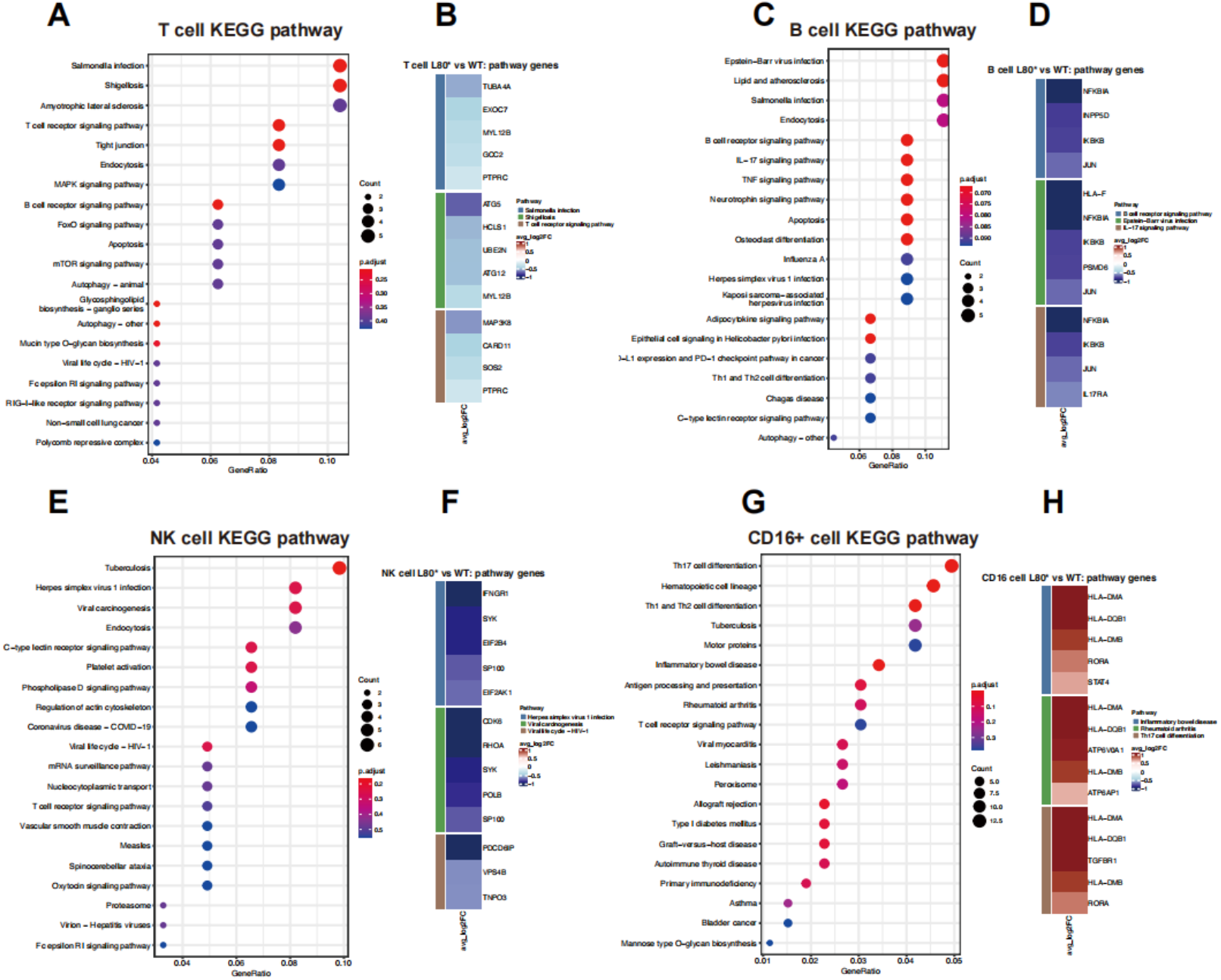
KEGG pathway enrichment analyses and pathway gene expression profiles in ELF4-mutant immune cell subsets. **(A, B)** KEGG pathway enrichment analysis **(A)** and corresponding heatmap of representative pathway genes **(B)** in T cells, comparing ELF4-mutant (L80*) and wild-type cells. **(C, D)** KEGG pathway enrichment analysis **(C)** and pathway gene expression heatmap **(D)** in B cells. **(E, F)** KEGG pathway enrichment analysis **(E)** and pathway gene expression heatmap **(F)** in NK cells, highlighting pathways related to antiviral responses and viral infection–associated signaling. **(G, H)** KEGG pathway enrichment analysis **(G)** and pathway gene expression heatmap **(H)** in CD16+monocytes, showing enrichment of inflammatory and immune-related pathways, including those associated with Th17 cell differentiation and inflammatory bowel disease.In dot plots, dot size represents the number of genes enriched in each pathway, and color indicates the adjusted P value. Heatmaps display the average log_2_fold change (avg_log_2_FC) of representative pathway genes comparing ELF4-mutant and wild-type cells within each cell type.

The most pronounced alterations were observed in NK cells and CD16^+^ monocytes. In NK cells, mutant populations showed marked downregulation of antiviral and interferon-signaling pathways (Fig. 3E, F), accompanied by reduced expression of key interferon-response genes such as *IFNGR1*. In contrast, ELF4-mutant CD16^+^ monocytes exhibited significant enrichment of pro-inflammatory pathways, including those involved in Th17 differentiation and inflammatory bowel disease (Fig. 3G, H). Taken together, these results demonstrate that somatic *ELF4* mutation drives cell-type specific pathway dysregulation, underscoring the broad role of ELF4 in immune homeostasis.

## Discussion

In this study, we describe a pediatric patient carrying a somatic truncating mutation in *ELF4*, who presented with recurrent mucocutaneous inflammation and periodic fever. Germline loss-of-function mutations in *ELF4* are known to cause DEX, a disorder characterized by heterogeneous inflammatory manifestations involving mucosal, gastrointestinal, and systemic features. Compared with previously reported germline cases^2–8^, the present patient exhibited a relatively restricted clinical phenotype. This difference likely stems from the somatic mosaic nature of the *ELF4* mutation, which limits the proportion of affected immune cells, rather than reflecting a less severe functional effect of the truncating variant itself. Supporting this interpretation, our functional analyses demonstrated that the ELF4 p.L80X variant leads to a clear loss-of-function at the molecular level, including markedly reduced protein expression.

By applying full length single-cell RNA sequencing, we directly compared ELF4-mutant and wild-type immune cells from the same individual, thereby assessing mutation-associated transcriptional programs while minimizing inter-individual variability. This approach^15^ revealed distinct transcriptional profiles in ELF4-mutant cells across both lymphoid and myeloid compartments, confirming that the somatic mutation drives transcriptional alterations in multiple immune lineages.

Notably, pathway-level analyses indicated that the impact of somatic ELF4 mutation is cell-type specific rather than uniform across the immune system. In adaptive immune cells such as T cells and B cells, altered pathways were primarily related to immune activation and lymphocyte signaling. The most pronounced pathway changes, however, were observed in innate immune subsets. In NK cells, ELF4-mutant populations showed downregulation of antiviral and interferon-related pathways^16^, along with reduced IFNGR1expression, suggesting impaired interferon responsiveness. Conversely, ELF4-mutant CD16^+^ monocytes exhibited enrichment of pro-inflammatory pathways linked to Th17 differentiation and inflammatory bowel disease, indicating a shift toward inflammatory transcriptional programs.

Collectively, these findings point to divergent immune biases associated with somatic ELF4 deficiency—namely, attenuated antiviral signaling in NK cells alongside heightened inflammatory activity in CD16^+^ monocytes. Rather than implicating a single pathogenic mechanism, this pattern suggests that partial disruption of ELF4 function differentially reshapes immune programs across cell types, contributing to autoinflammatory disease in a context-dependent manner.

## Data Availability

All data produced in the present study are available upon reasonable request to the authors

## Acknowledgement

We sincerely thank the patient and his family for their participation in this study. We thank Dr. Yu Liang (Department of Pediatric Nephrology and Rheumatology, The Third Affiliated Hospital of Zhengzhou University) for his assistance in communicating with the patient’s family.

## Authorship contribution statement

**Qianlu Zhang:** Conceptualization, Methodology, Formal analysis, Writing - Original Draft. **Yatang Lei:** Data curation, Writing - Original Draft. **Xiaodong Zhao:** Supervision, Project administration, Review & Editing, Funding acquisition. **Hongqiang Du:** Conceptualization, Supervision, Writing - Review & Editing, Funding acquisition.

## Ethics declaration

This study was conducted in accordance with the Declaration of Helsinki and was approved by the Institutional Review Board of the Children’ s Hospital of Chongqing Medical University (Approval No. 2021-138). Written informed consent was obtained from the parents of all participants prior to inclusion in the study.

## Data availability

The data that support the findings of this study are available from the corresponding authors upon reasonable request.

